# The Chinese Assessment of physical literacy:Based on Grounded Theory Paradigm for children in grades 3-6

**DOI:** 10.1101/2022.01.12.22269057

**Authors:** Wang YongKang, Fu QianQian

## Abstract

The aim of this study is to construct and validate “physical literacy self-assessment questionnaire”(PLAQ)for Chinese students in grades 3-6. This study uses qualitative and quantitative methods to construct evaluation indicators of PL and determine the weights of each indicator. The 60 items of original PLAQ was based on literature review and interviews,and administered to 1179 primary students graded 3-6 in China. Exploratory factor analysis(EFA)and confirmatory factor analysis(CFA) are used to optimize the structure and verify the reliability and validity of the questionnaire. The model of PLAQ is composed of 4 first-level indicators, 10 second-level indicators and 35 third-level indicators. The results of EFA and CFA resulted in a 44-items, 4-factor questionnaire. EFA item loadings ranged from 0.558 to 0.896,and Cronbach’s alpha ranged from 0.818 to 0.892. The results of CFA show that the constructed model fits well, and PLAQ has good convergent validity and discriminative validity. The PLAQ appeared to be reliable and valid that can be used as an assessment tool for students in grades 3-6. PLAQ can be used as a guide for the development of PL. Additionally, PLAQ gives us a shared understanding about what PL is and how it can be developed by Chinese children. However, studies on the accuracy and generalizability of the PLAQ should be conducted to improve it in the future.

## 1. Introduction

The sedentary lifestyle has become a major health problem for human. Of the 56 million people who die each year, 3.2 million deaths (6 deaths per minute) can be attributed to lack of physical activity [1]. A survey in Australia shows that declining physical activity and rising chronic diseases are driving up public health costs [2]. When the “health-based models” is not enough to change people’s sedentary lifestyle, Whitehead proposes the concept of physical literacy(PL) based on monism, phenomenology and existentialism. PL tries to promote the participation of physical activity in the whole process of life. As soon as the concept of PL was put forward, it had a great impact and gradually became an important concept to solve the global physical inactivity.

The concept of PL was proposed by Whitehead in 1993 and has been refining it ever since. For example,in 2002 PL was defined as “The characteristics of a physically literate individual are that the person moves with poise, economy and confidence in a wide variety of physically challenging situations. In addition the individual is perceptive in ‘reading’ all aspects of the physical environment, anticipating movement needs or possibilities and responding appropriately to these, with intelligence and imagination”[3]. In 2010,PL was described as”The motivation, confidence, physical competence, knowledge, and understanding to maintain physical activity throughout the lifecourse”[4]. In 2013,Whitehead had described physical literacy in the International Council for Sport Science and Physical Education (ICSSPE) bulletin as “the motivation, confidence, physical competence, knowledge, and understanding to value and take responsibility for maintaining purposeful physical pursuits/activities throughout the lifecourse”[5][6]. Based on Whitehead’s definition of PL, scholars from different countries put forward the concept of localized PL in combination with regional culture. The Canadian Healthy Active Living and Obesity Research Group(HALO)and Canada Sports for Life(CS4L) are consistent with IPLA that PL is the motivation, confidence, physical competence, knowledge, and understanding to value and take responsibility for engagement in physical activities for life[7][8]. Physical and Health Education Canada(PHE Canada) definition is:”moving with competence and confidence in a wide variety of physical activities in multiple environments that benefit the healthy development of the whole person”[9]. Sport Australia(SA)defined PL is about developing the skills, knowledge and behavior that give us the confidence and motivation to lead active lives. It can help people at every stage of life develop and maintain positive physical activity behavior and delivers physical, psychological, social and cognitive health and wellbeing benefits [1][10]. The concept of physical literacy presents a pluralistic feature in the world. The diversity of physical literacy concepts does not mean chaos and disorder, but shows its strong vitality and promotes the development of physical literacy in the world.

Different cultures, governance structures, geographical locations and physical environments may lead to the need to construct diversified PL assessment tools and promotion strategies [11]. Three assessments and evaluations of PL have been established and are used widely [12]: the Canadian Assessment of Physical Literacy (CAPL)[13];Physical Literacy Assessment for Adolescents(PLAY);and the Passport for Life (P4L). Although there are differences in the target population, evaluation methods, evaluation indicators and scoring standards of the three tools, different research perspectives further promote the development of the theoretical system of physical literacy.

With more and more countries take PL as the operation guide of physical education and physical activity curriculum design in primary and secondary schools. A heated discussion on PL is expanding in the field of international physical education. The United States takes PL as the objective and guiding ideology of physical education in primary and secondary schools [14]. Canada combines the concept of PL with physical education, federal education regulations and long-term planning for athletes [15]. In 2013, the British began to promote “Primary School Physical Literacy Framework”, is aimed at providing support for developing each elementary student’s PL. The framework proposed “PL is not a plan, but the result of organized physical education and competitive sports” [14].

Over the past few decades, nearsightedness and obesity have increased among Chinese children and adolescents due to lack of physical activity. In order to improve the health problems faced by Chinese children and adolescents,some specific measures were taken that include increasing the number of school PE courses, reducing homework, increasing after-school physical activity time, developing students’ interest in sports and sports skills. In 2019,the new plan named “Outline for Building a Leading Sports Power”, which clearly stated that “promoting children and adolescents to improve their physical literacy and develop a healthy lifestyle will be an important part of school physical education” [16]. Policy makers try to promote these goals by adopting the idea of physical literacy. Some scholars believe that physical literacy can play a fundamental role only when entering schools, especially primary schools,which has a great influence on the subsequent state of physical literacy [17][18]. Due to China’s vast territory and large primary school students, it is necessary to develop a self-assessment tool for measuring the physical literacy of Chinese primary school students in order to assess their own PL level quickly and accurately.

## 2. Methods

### 2.1 Paradigm of grounded theory process to develop the PL evaluation indicators

#### 2.1.1 Participants

The approval for interview on human subjects was obtained from the Academic Committee of the School of Physical Education and Sport Science, Fujian Normal University. Grounded theory requires entering the world of research objects, seeing their world from the perspective of research objects, and understanding their views and behaviors [19]. Therefore, primary school students, parents, teachers and experts were selected as the interviewees in this study. In this study, 32 PE teachers in primary schools, 30 primary school students, 20 parents of students and 8 experts and scholars in this field were selected as the interviewees from Beijing, Tianjin, Shanxi, Shaanxi, Hebei, Jiangsu and Zhejiang provinces and municipalities. The topics of the interview include the knowledge of physical literacy, the influencing factors of physical literacy, and the situation of participation in physical activity. The interviewee is informed in advance of the research theme and the main content of the interview, and first-hand information is obtained through semi-structured interviews. After the interview, the recording files were converted into text materials, and the interview data was coded and sorted through Nvivo 12.0.

#### 2.1.2 Coding Process

##### 2.1.2.1 Open coding

In the open coding stage, the original interview data is coded paragraph by paragraph, sentence by sentence, line by line, and word by word to obtain the initial concept and initial category. In the open coding process, in order to reduce the impact of the coder’s academic background, subjective cognition, emotional fluctuations, etc. on the coding results, the coders try to extract the original words or sentences used by the interviewee as the initial concept.

##### 2.1.2.2 Axial Coding

The main category is extracted by merging the initial categories of the same concept connotation, and the initial category is the merging of the initial concepts with the same connotation. Therefore, the initial concept, the initial category and the main category are related to each other. Through constant comparison of concepts, this study extracts the multi-domain structure of physical literacy from the data, including 4 main categories, 10 sub-categories and 35 concepts. See Table 3 for details.

##### 2.1.2.3 Selective coding

Selective coding is to explore the relationship between coding and coding, concept and concept, category and category, link these relationships in a certain way, and outline the preliminary presented theory. Further determine the connotation and extension of the theory, return the theory to the original data for verification, and continuously optimize the existing theory to make it more refined.

#### 2.1.3 Using AHP to determine weights of the evaluation indicators

Analytical Hierarchy Process (AHP) is used to calculate the weights of physical literacy evaluation indicators, 12 experts in the field of physical literacy and 8 primary physical education teachers with senior titles are selected to judge the importance of physical literacy evaluation indicators. Using Yaahp 10.1 software carry out the eights of physical literacy evaluation indicators (see Table 5).

### 2.2 The verification process of the self-assessment questionnaire on physical literacy

#### 2.2.1 Designing the Physical literacy self-assessment questionnaire(PLAQ)

Compile original PLAQ items according to the evaluation indicators of physical literacy. The original PLAQ consists of 60 items scored on a 5-point Likert scale (1 =strongly disagree and 5 = strongly agree) (see Table 2).The 60 items of the original PLAQ are equally divided into four subscales including: “physical competence”(13 items),”affective domain(” 16 items),”knowledge and understanding” (15 items), and “behavior of physical activity” (15 items). The questionnaire items were tested for content validity through expert interviews.

**Table 1.** Original 60-items of PLAQ for Chinese 3-6 grades students Sign Items

| Sign | Items |
| --- | --- |
| A1 | I am satisfied with my muscle strength. |
| A2 | I can run fast. |
| A3 | I am good at throwing or other strength sports. |
| A4 | When it comes to long-distance running, I am better than most of my peers. |
| A5 | Compared with my peers, my body movements are not very coordinated. |
| A6 | I can stretch and rotate my body well. |
| A7 | When I exercise, my movements are graceful and coordinated. |
| A8 | I can master basic movements such as walking, running and jumping better than my peers. |
| A9 | I can master the basic motor skill of running, throwing, kicking, catching and jumping. |
| A10 | I am good at coordinated (consistent) movements |
| A11 | I'm good at sports. |
| A12 | Most sports are easy for me. |
| A13 | I am better at sports than most of my friends. |
| B1 | I often want to participate in physical activity. |
| B2 | I am willing to try new sports. |
| B3 | I know what sports I am good at. |
| B4 | I am willing to communicate and express ideas with my peers. |
| B5 | Show tenacity and self-confidence after experiencing setbacks. |
| B6 | Full of confidence when learning sport skills. |
| B7 | Believe that you do better than others during exercise. |
| B8 | Congratulations to the teammates for their good performance in sports. |
| B9 | I will not blame, criticize, or abuse my teammates. |
| B10 | Malicious foul, trying to hurt an opponent. |
| B11 | Request to stop the game or competition when the opponent is injured |
| B12 | Have a spirit of fair competition when participating in sports or games. |
| B13 | Have the spirit of teamwork when participating in sports or games. |
| B14 | Have a tenacious spirit when participating in sports activities or games. |
| B15 | I can resolve my bad emotions through exercise. |
| B16 | I can control the bad emotions during exercise. |
| B17 | Once the body is uncomfortable ( fatigue, pain, nausea, etc. ) during exercise, I can make adjustments in time. |
| C1 | I know the names of a lot of sports. ( For example : football , basketball , swim ) |
| C2 | I know the names of many sport skill. ( For example :dribble ;one hand shoulder shot ) |
| C3 | I am familiar with sports tactics. |
| C4 | I am familiar with the rules of many sports. |
| C5 | Do not swim in dangerous waters. |
| C6 | Warm up before physical activity and relax after physical activity. |
| C7 | When the air is bad, I usually avoid outdoor physical activities. |
| C8 | I don't exercise vigorously immediately after a meal. |
| C9 | I know how to drink water during exercise. |
| C10 | When learning difficult or dangerous techniques, I usually do it under the guidance of a teacher or coach. |
| C11 | If I feel uncomfortable during physical activities, I will seek help from teachers, parents and doctors. |
| C12 | I know the dangers of sedentary. |
| C13 | I know the advantages of physical activity. |
| C14 | I try to avoid high-calorie foods. |
| C15 | I know how to improve physical activity. |
| D1 | I spend more than 2 hours on the electronic screen every day. |
| D2 | I participate in sports activities (including sports classes and extracurricular activities) and games no less than 5 times a week. |
| D3 | Sports activities (including physical education classes and extracurricular activities) are not less than 1 hour per day. |
| D4 | Without affecting my study and life, I usually choose to travel on foot or by bike. |
| D | After physical activity, I sleep well. |
| D6 | I can exercise in different seasons. |
| D7 | I can exercise on snow, water, and on land. |
| D8 | After changing coaches or teachers, adapt to their teaching methods faster than their peers. |
| D9 | When I encounter difficulties in physical activity, I will not give up easily and seek solutions. |
| D10 | I maintain moderate to high intensity physical activity. |
| D11 | Regardless of whether my parents support me to actively participate in physical activities, I maintain my love for sports. |
| D12 | Regardless of my relationship with the teacher, it does not affect my exercise and learning. |
| D13 | When sports facilities are insufficient, I can find a solution. |
| D14 | Often create new rules of the game with my companions. |
| D15 | I often take the initiative to participate in physical activities. |

**Table2.**
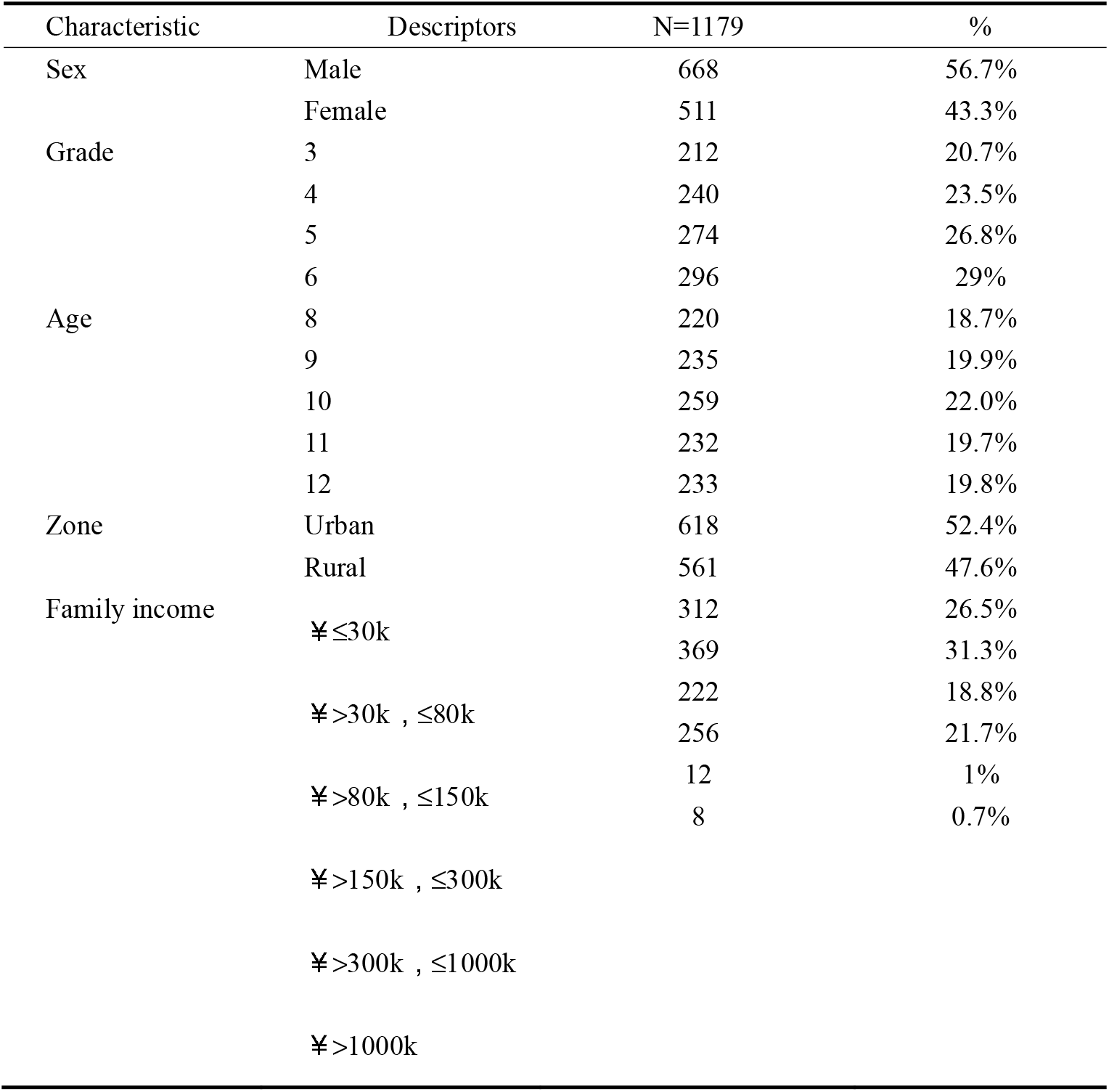
Children’ demographic characteristics

#### 2.2.2 Data collection and sampling

The experiment was approved by Academic Committee of the School of Physical Education and Sport Science, Fujian Normal University. The participants of this survey were Chinese children from primary schools in China. A total of 1179 Chinese children from 8 randomly selected primary schools (grade 3-6) agreed to participate in this study after letters of invitation were sent to their parents,school principals and PE teachers. All participants were asked to provide informed consent with emphasis on the voluntary nature of the survey before participating in the study.

#### 2.2.3 Data analysis and results

The researcher used SPSS version 21 for Windows for the data analysis. In advance of validation of the PLAQ, children’ demographic characteristics (gender, grade,zone and family income) were analyzed in Table 2. After confirming the validity of the data, the researchers conducted exploratory factor analysis(EFA)and confirmatory factor analysis(CFA).

#### 2.2.4 Exploratory factor analysis(EFA)

In the process of constructing the questionnaire for each domain, in order to verify whether the variable is suitable for factor analysis, Kaiser-Mayer-Olkin (KMO) and Bartlett sphere tests should be performed first for all items of the domain, if KMO≥ 0.7, and Bartlett sphere test result is significant (p<0.05), which indicates that the variable of this domain is suitable for factor analysis [20].

In the process of constructing the questionnaire in this study, most of the questionnaire items used the Likert-5 point method, and all variables were non-continuous variables. The number of extracted factors is judged according to the Kaiser standard and lithographs, and the feature value of the extracted factors >1 meets the requirements of the inflection point of lithographs. In the theoretical construction, the sub-domains in the core domain are not completely independent of each other, and there is a certain correlation between the sub-domains. Therefore, the factor rotation adopts the oblique rotation method of Promax. The criteria for item exclusion are as follows [21]:

- Items with too small factor loading, factor loading<0.5;
- The load of the item in multiple factors>0.5;
- For items whose cross loads are too close, the difference between the absolute values of the cross loads of the items is<0.2.

EFA is carried out on the four core domains of physical literacy. Each domain may need to go through multiple rounds to determine the results of the final item screening. The research results are only presented in detail for the final EFA.

#### 2.2.5 Confirmatory factor analysis (CFA)

CFA is to evaluate the model of each domain of physical literacy on the basis of EFA, and to screen the problematic items again. In order to ensure a good fit of the overall model and a distinction between various latent variables, the overall CFA model needs to meet the following requirements [22]:

- NC(CMIN/DF) <3;
- RMSEA<0.8;
- CFI>0.9;
- TLI>0.9;
- IFI>0.9;
- GFI>0.9.

The measurement model needs to meet the following requirements:

- Entry factor loading>0.45

In confirmatory factor analysis, if the overall fit of the model is not high, the model will be revised. In order to ensure the independence of each latent variable, the revision of the model only increases the residual correlation of items under the latent variable. In the measurement model, if an entry does not meet the indicator requirements of the measurement model, this entry will be deleted, the CFA of the entire model will be performed again, and the model results obtained will be re-verified.

## 3. Results

### 3.1 Evaluation indicators of Physical literacy for Chinese primary students based on Grounded Theory

After open coding the interview records of all interviewees, the following open coded results were obtained, and 272 initial concepts were obtained. Through further comparison, it was found that some of the 272 initial concepts were repeated, and 35 non-repeated initial concepts were finally obtained.

The main category is extracted by merging the initial categories of the same concept connotation, and the initial category is the merging of the initial concepts with the same connotation. Therefore, the initial concept, the initial category and the main category are related to each other. And in the process of inducing similar items, concepts or initial categories that have two or more adjacent connotations must be satisfied before they can be classified. Therefore, the main category has the following characteristics:

- Core characteristics that can be correlated with other data;
- Can explain most of the behavior of the research topic;
- Frequent reproducibility;
- Relevant and meaningful with other variables.

Based on the above principles and through constant comparison of concepts, the axial coding results show the multi-dimensional structure of PL, including 4 main categories, 10 sub-categories and 35 concepts, including physical ability, motivation and confidence, sports moral, self-regulation, knowledge of sports safety and risk, basic knowledge of sports, knowledge of physical activity value and exercise promotion,physical activity level and adaptive capacity(see Table 4).

**Table 3.** Open coding results (example)

| Initial category | Initial concept | Original sentence |
| --- | --- | --- |
| Take an exam | Exercise in order to pass the physical exam. | Some children told me that they actively exercise in order to pass the physical examination. ( T ) |
| Experience | Children cannot | Now many children have no experience. They don't |
|  | perceive the benefits without experiencing sports. | know the benefits. Sports are to be experienced. If you don't cross the horizontal bar, you will not feel the joy of success. If you haven't competed, you can't experience the joy of the game. (E) |
| Family financial support | Parents provide children with funds for exercise. | I support my child to exercise, and will take child to apply for some sport training courses, these things can promote my child to develop good habits of exercise. (P) |
| Teaching method | Inappropriate teaching methods. | Some teachers' teaching methods are rather rigid. They run laps when warming up in class and don't play games, which causes students to lose the fun of exercise. (E) |
| Confidence | Courage to try. | I like to try all kinds of games that I haven't played. (S) |

*NOTE :T= result of teacher interview ;E= result of expert interview ;P= result of parent interview ; S= result of student interview.*

**Table 4.**
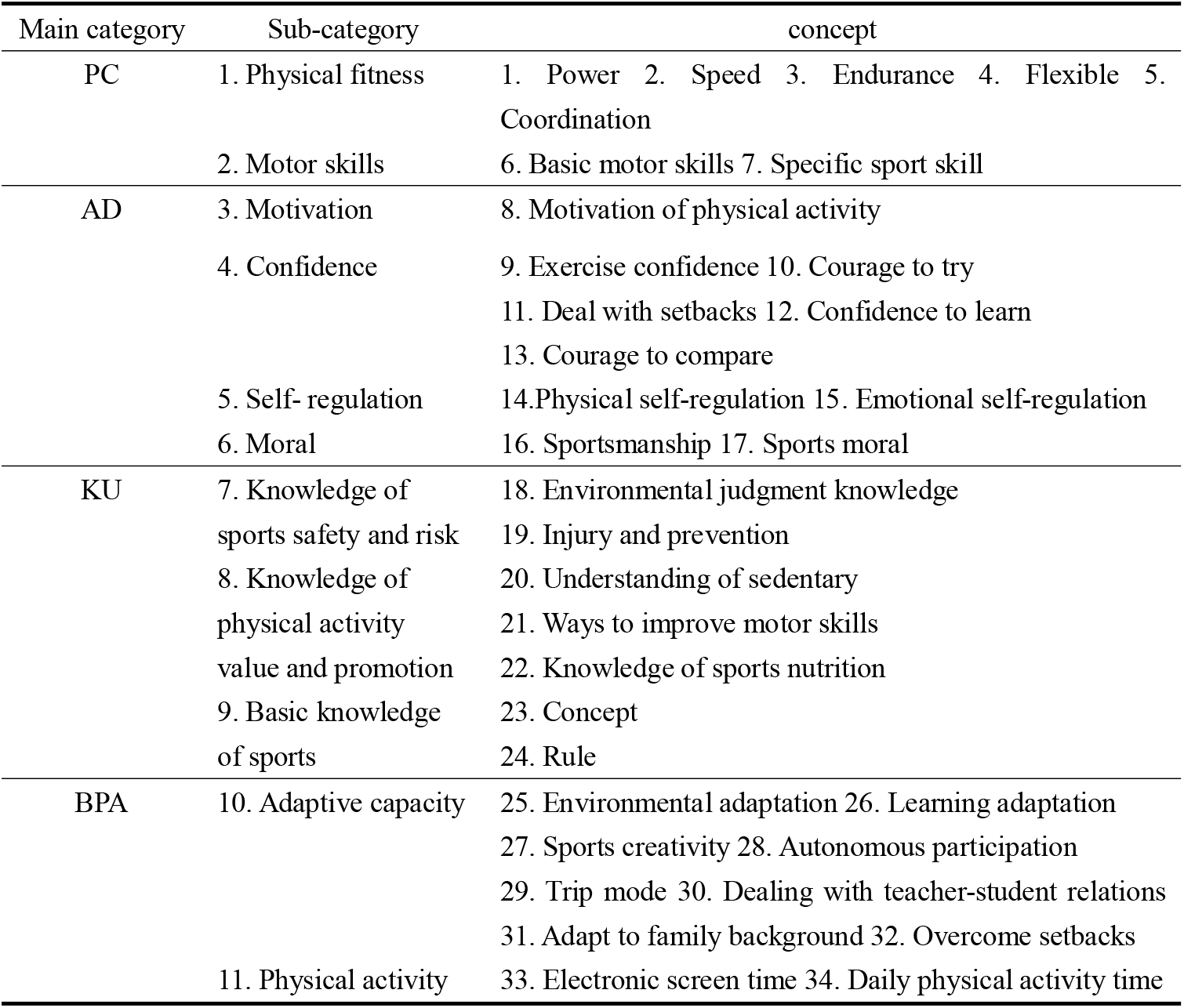
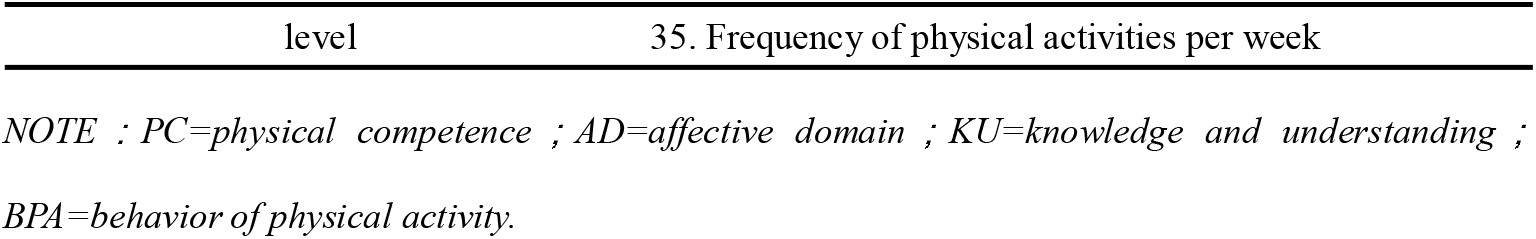
Axial coding results

Selective coding is a process to further explore the relationship between main category and main category, sub-category and sub-category, and concept and concept, and form the structure model of PL. Through the continuous comparison of concepts and the combination of interview data, the study believes that the four attributes (physical competence,affective,Knowledge and understanding,Physical activity behavior)of PL influence each other(see Figure 2).

**Figure1.**
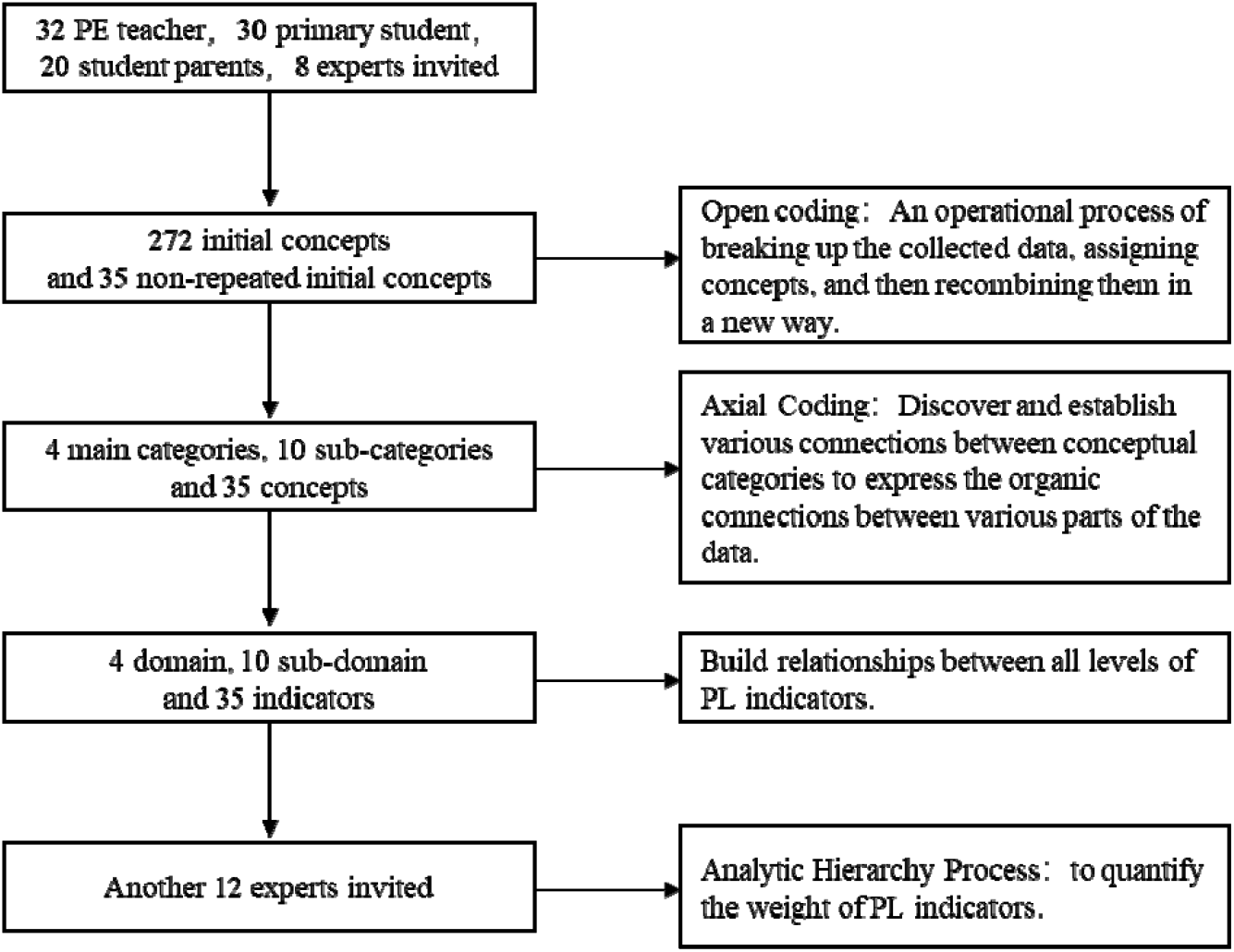
Paradigm of grounded theory process.

**Figure 2.**
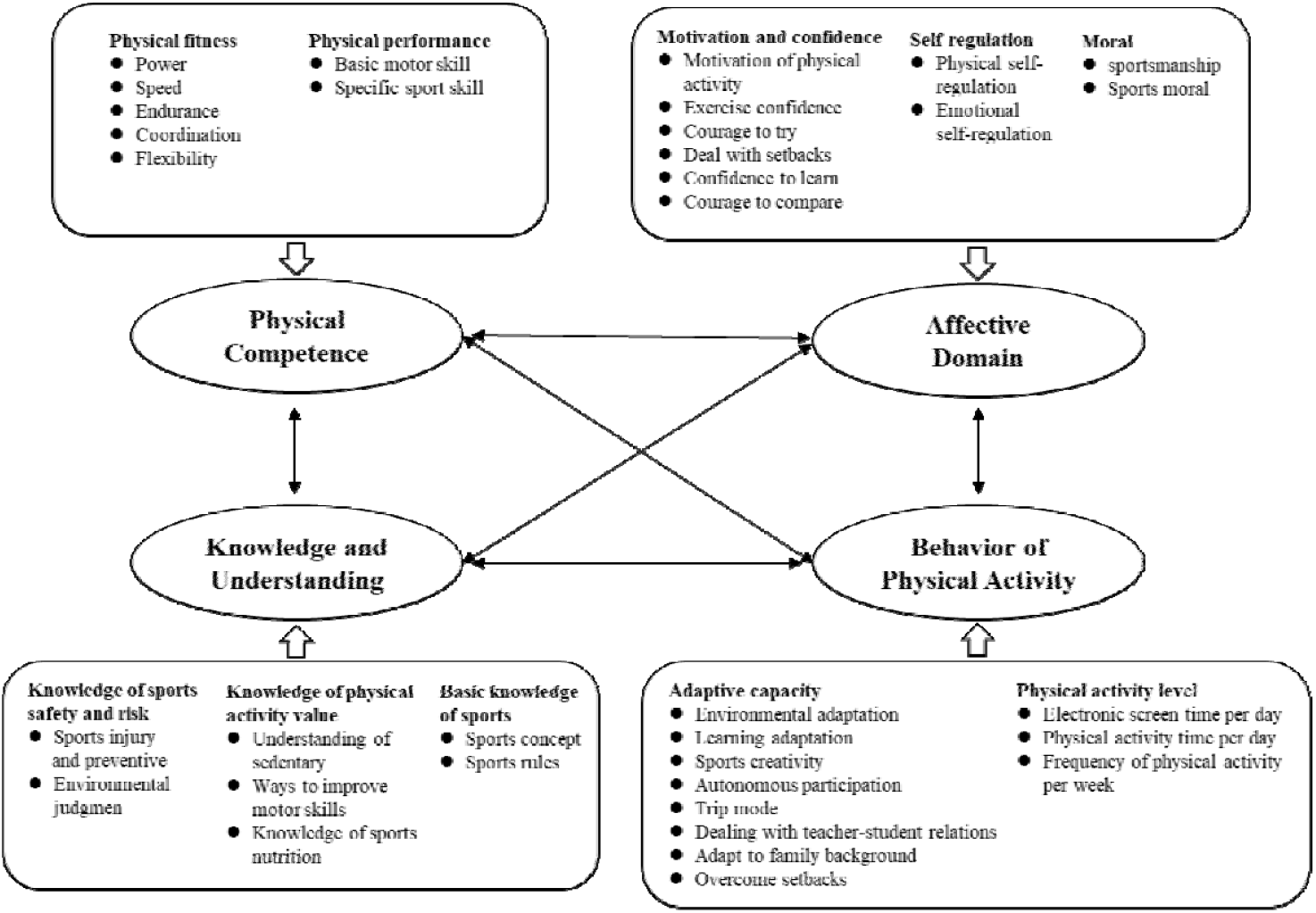
The physical literacy structure model for Chinese students in grade 3-6.

The theoretical saturation determines the sample size. In order to verify the theoretical saturation of the selected sample, after all the data coding was completed, additional interviews were conducted with 5 PE teachers,5 primary students,5 parents and 3 experts. After coding, no new concept was formed, that is, there was no new category and core categories indicates that have reached theoretical saturation.

It can be seen from Table 5 that in the first-level indicators of PL, the weights of physical competence, affective domain, knowledge and understanding, and behavior of physical activity are respectively 20.48%, 28.81%, 16.90%, and 33.81% (see Table 5).

**Table 5.** The weights of the physical literacy evaluation indicators for Chinese students in grade 3-6

| First level indicator | Secondary indicators | Three-level indicators |
| --- | --- | --- |
| PC ( W=0.2048 ) | Physical fitness ( W=0.0683 ) | Power ( W=0.0130 ) |
|  |  | Speed ( W=0.0094 ) |
|  |  | Endurance ( W=0.0052 ) |
|  | Physical performance<br>( W=0.1365 ) | Coordination ( W=0.0212 ) |
|  |  | Flexibility ( W=0.0195 ) |
|  |  | Basic motor skill ( W=0.0683 ) |
|  |  | Specific sport skill ( W=0.0683 ) |
| AD ( W=0.2881 ) | Motivation and confidence | Motivation of physical activity |
|  | ( W=0.1198 ) | ( W=0.0311 ) |
|  |  | Exercise confidence ( W=0.0145 ) |
|  |  | Courage to try ( W=0.0136 ) |
|  |  | Deal with setbacks ( W=0.0333 ) |
|  |  | Confidence to learn ( W=0.0128 ) |
|  | Self-regulation ( W=0.0364 ) | Courage to compare ( W=0.0145 ) |
|  |  | physical self- regulation |
|  |  | ( W=0.0182 ) |
|  | Moral ( W=0.1319 ) | Emotional self- regulation |
|  |  | ( W=0.0182 ) |
|  |  | sportsmanship ( W=0.0659 ) |
|  |  | Sports moral ( W=0.0659 ) |
| KU ( W=0.1690 ) | Knowledge of sports safety and | Sports injury and preventive |
|  | risk ( W=0.0960 ) | ( W=0.0480 ) |
|  |  | Environmental judgment |
|  |  | ( W=0.0480 ) |
|  | Knowledge of physical activity | Understanding of sedentary |
|  | value and promotion ( W=0.0565 ) | ( W=0.0226 ) |
|  |  | Ways to improve motor skills |
|  |  | ( W=0.0226 ) |
|  | Basic knowledge of sports | Knowledge of sports nutrition |
|  | ( W=0.0166 ) | ( W=0.0113 ) |
|  |  | Sports concept ( W=0.0083 ) |
|  |  | Sports rules ( W=0.0083 ) |
| BPA ( W=0.3381 ) | Adaptive capacity ( W=0.2536 ) | Environmental adaptation<br>( W=0.0519 ) |
|  |  | Learning adaptation ( W=0.0164 ) |
|  |  | Sports creativity ( W=0.0172 ) |
|  |  | Autonomous participation<br>( W=0.0597 ) |
|  |  | Trip mode ( W=0.0281 ) |
|  |  | Dealing with teacher-student<br>relations ( W=0.0178 ) |
|  |  | Adapt to family background<br>( W=0.0205 ) |
|  |  | Overcome setbacks ( W=0.0418 ) |
|  |  | Electronic screen time per day<br>( W=0.0282 ) |
|  |  | Physical activity time per day<br>( W=0.0282 ) |
|  |  | Frequency of physical activity per<br>week ( W=0.0282 ) |

### 3.2 Verification of PLAQ for Chinese 3-6 grades students

#### 3.2.1 Bartlett’s Test of Sphericity and KMO index

Table 6 shows the results of the Bartlett’s Test of Sphericity and KMO test for the core domains of physical literacy. From the results of the Bartlett’s Test of Sphericity, p<0.01 should reject the null hypothesis, indicating that the four core domains have common factors. From the KMO test value, the KMO value of the four core domains is between 0.8-0.9. When KMO > 0.8, it indicates that it is suitable for factor analysis [23].

**Table 6.** Fitness test of factor analysis of physical literacy sub-questionnaire

| Core domains | Bartlett |  |  | KMO |
| --- | --- | --- | --- | --- |
| | $\chi^2$ | df | p | |
| PC | 1019.342 | 171 | <0.01 | 0.818 |
| AD | 2075.966 | 300 | <0.01 | 0.892 |
| KU | 1505.528 | 210 | <0.01 | 0.885 |
| BPA | 952.904 | 120 | <0.01 | 0.891 |

NOTE : PC=physical competence ; AD=affective domain ; KU=knowledge and understanding ; BPA=behavior of physical activity

#### 3.2.2 Results of Exploratory Factor Analysis (EFA)

In order to test the validity of the questionnaire, a factor analysis of the questionnaire is needed. The purpose is to explore the potential structure of the questionnaire and reduce the number of items. Studies have shown that if the questionnaire is prepared by clearly dividing the scale into several subscales (levels or constructs) based on references and related theories, the item variables included in each subscale should be used for factor analysis separately, instead of the whole scale for factor analysis [24].

First, the Bartlett sphere test and KMO test are used to calculate whether the four sub-questions are suitable for factor analysis. The research results show that the KMO values of the four sub-questionnaires of PC, AD, KU, and BPA are 0.818, 0.892, 0.885, and 0.891, and the chi-square value is 1019.342 (df=171, p<0.01), 2075.966 (df=300, p<0.01), 1505.528 (df=210, p<0.01), 952.904 (df=120, p<0.01). When KMO>0.7, it is suitable for factor analysis. The KMO value of the four core domains is between 0.8-0.9, therefore, the four sub-questionnaires are suitable for factor analysis.

Using the principal component analysis method, items with low factor loading or cross-loading were deleted, and 16 items were finally deleted. The PC sub-questionnaire deletes four items A3, A7, A10 and A12(see Table 7). The AD sub-questionnaire deletes four items B4, B8, B10 and B11(see Table 8). The KU sub-questionnaire deletes four items C3, C8, C9 and C12(see Table 9). The BPA sub-questionnaire deletes four items D5, D6, D7 and D10(see Table 10). There are a total of 10 factors with factor eigenvalues greater than 1, and 44 items are retained.

**Table 7.**
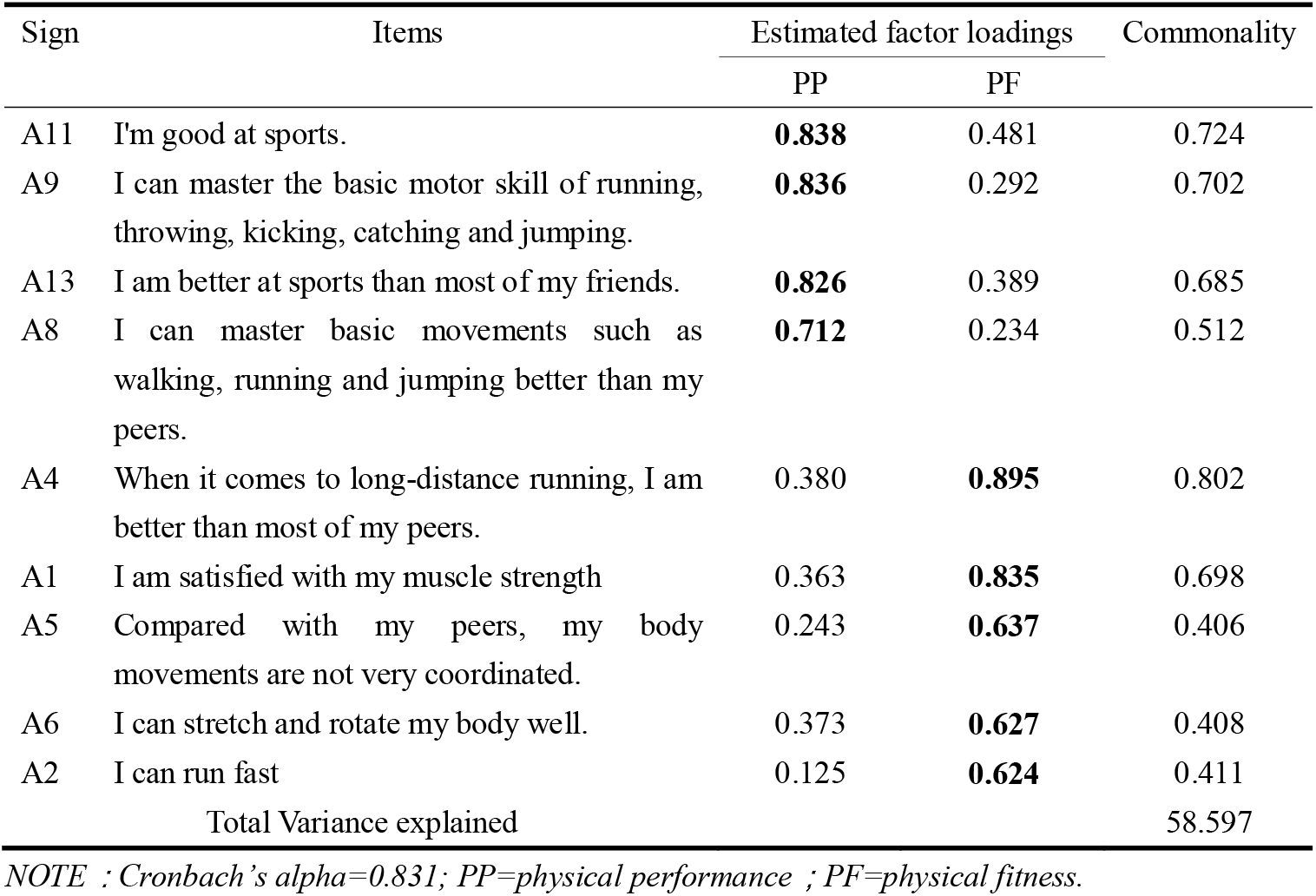
Factor structures of physical competence by exploratory factor analysis

**Table 8.** Factor structures of affective domain by exploratory factor analysis

| Sign | Items | Estimated factor loadings |  |  | Commonality |
| --- | --- | --- | --- | --- | --- |
|  |  | MC | SR | MOR |  |
| B5 | Show tenacity and self-confidence after experiencing setbacks. | **0.811** | 0.333 | 0.161 | 0.793 |
| B6 | Full of confidence when learning sport skills. | **0.797** | 0.312 | 0.091 | 0.741 |
| B7 | Believe that you do better than others during exercise. | **0.787** | 0.286 | -0.025 | 0.702 |
| B2 | I am willing to try new sports. | **0.779** | 0.088 | 0.307 | 0.709 |
| B3 | I know what sports I am good at. | **0.736** | 0.366 | 0.018 | 0.676 |
| B1 | I often want to participate in physical activity. | **0.531** | 0.153 | 0.253 | 0.370 |
| B17 | Once the body is uncomfortable ( fatigue, pain, nausea, etc. ) during exercise, I can make adjustments in time. | 0.155 | **0.796** | 0.081 | 0.664 |
| B16 | I can control the bad emotions during exercise. | 0.286 | **0.792** | 0.115 | 0.722 |
| B15 | I can resolve my bad emotions through exercise. | 0.381 | **0.736** | 0.178 | 0.718 |
| B12 | Have a spirit of fair competition when participating in sports or games. | 0.438 | 0.313 | **0.671** | 0.740 |
| B9 | I will not blame, criticize, or abuse my teammates. | 0.471 | 0.243 | **0.611** | 0.653 |
| B13 | Have the spirit of teamwork when participating in sports or games. | 0.492 | 0.358 | **0.601** | 0.732 |
| B14 | Have a tenacious spirit when participating in sports activities or games. | 0.501 | 0.426 | **0.558** | 0.743 |
| Total Variance explained |  |  |  |  | 67.866 |

NOTE :Cronbach's alpha=0.908; MC=motivation and confidence ;SR=self- regulation ;MOR=moral.

**Table 9.**
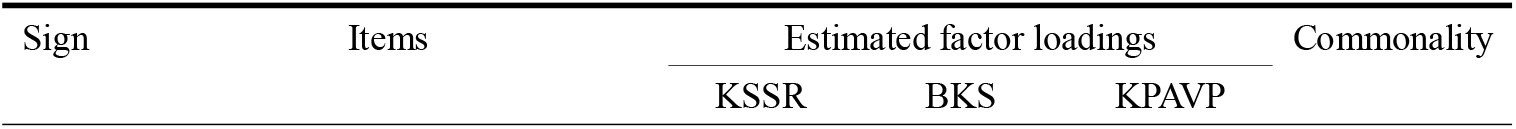
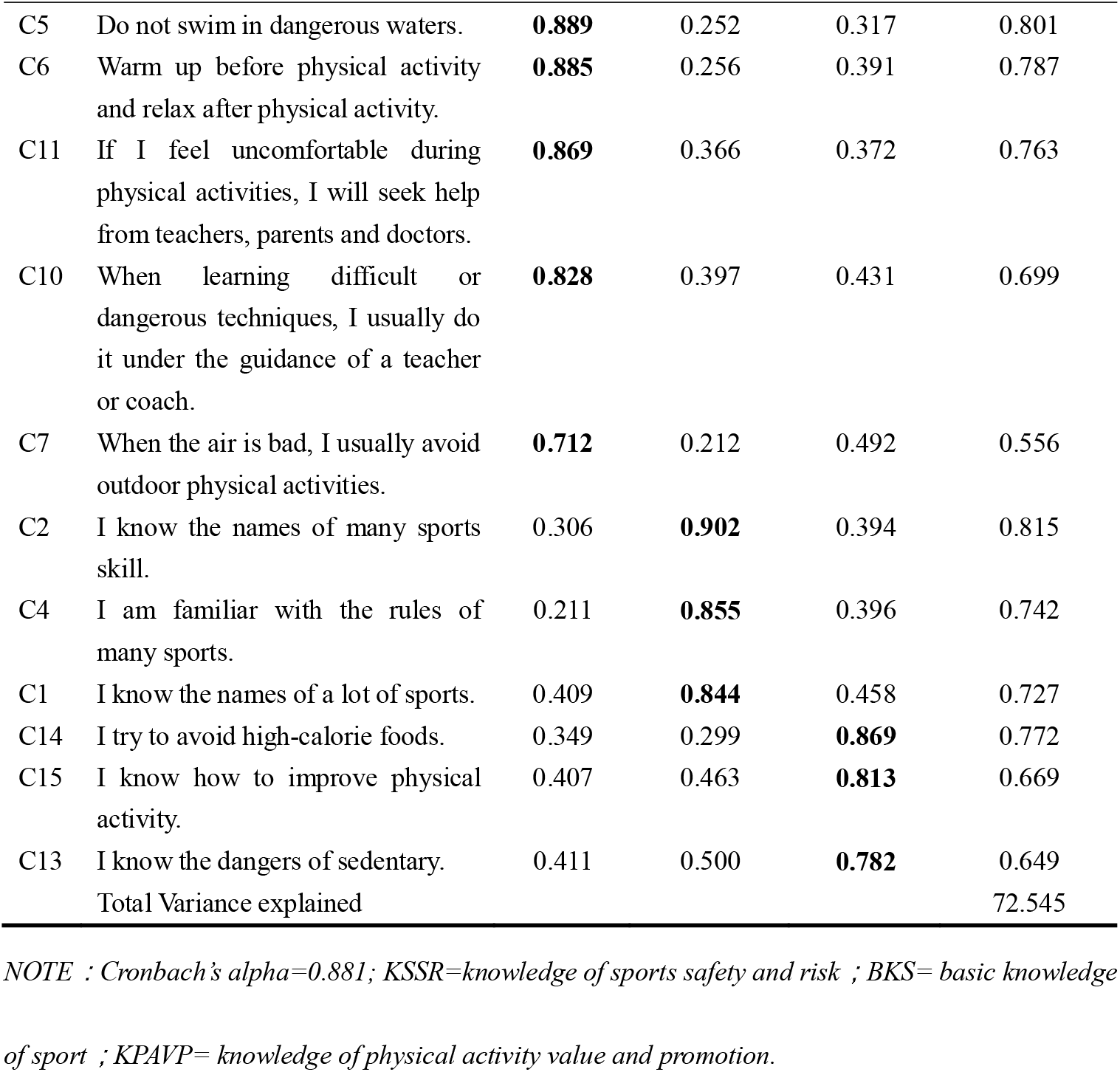
Factor structures of knowledge and understanding by Exploratory Factor Analysis

| Sign | Items | Estimated factor loadings |  |  | Commonality |
| --- | --- | --- | --- | --- | --- |
|  |  | KSSR | BKS | KPAVP |  |
| C5 | Do not swim in dangerous waters. | **0.889** | 0.252 | 0.317 | 0.801 |
| C6 | Warm up before physical activity and relax after physical activity. | **0.885** | 0.256 | 0.391 | 0.787 |
| C11 | If I feel uncomfortable during physical activities, I will seek help from teachers, parents and doctors. | **0.869** | 0.366 | 0.372 | 0.763 |
| C10 | When learning difficult or dangerous techniques, I usually do it under the guidance of a teacher or coach. | **0.828** | 0.397 | 0.431 | 0.699 |
| C7 | When the air is bad, I usually avoid outdoor physical activities. | **0.712** | 0.212 | 0.492 | 0.556 |
| C2 | I know the names of many sports skill. | 0.306 | **0.902** | 0.394 | 0.815 |
| C4 | I am familiar with the rules of many sports. | 0.211 | **0.855** | 0.396 | 0.742 |
| C1 | I know the names of a lot of sports. | 0.409 | **0.844** | 0.458 | 0.727 |
| C14 | I try to avoid high-calorie foods. | 0.349 | 0.299 | **0.869** | 0.772 |
| C15 | I know how to improve physical activity. | 0.407 | 0.463 | **0.813** | 0.669 |
| C13 | I know the dangers of sedentary. | 0.411 | 0.500 | **0.782** | 0.649 |
| Total Variance explained |  |  |  |  | 72.545 |

NOTE : Cronbach's $\alpha=0.881$ ; KSSR=knowledge of sports safety and risk ; BKS= basic knowledge of sport ; KPAVP= knowledge of physical activity value and promotion.

**Table 10.** Factor structures of behavior of physical activity by Exploratory Factor Analysis

| Sign | Items | Estimated factor loadings |  | Commonality |
| --- | --- | --- | --- | --- |
|  |  | AC | PAL |  |
| D12 | Regardless of my relationship with the teacher, it does not affect my exercise and learning. | **0.896** | 0.249 | 0.827 |
| D9 | When I encounter difficulties in physical activity, I will not give up easily and seek solutions. | **0.887** | 0.339 | 0.789 |
| D11 | Regardless of whether my parents support me to actively participate in physical activities, I maintain my love for sports. | **0.860** | 0.214 | 0.772 |
| D15 | I often take the initiative to participate in physical activities. | **0.811** | 0.335 | 0.658 |
| D4 | I usually choose to travel on foot or by bike. | **0.768** | 0.420 | 0.632 |
| D13 | When sports facilities are insufficient, I can find a solution. | **0.762** | 0.491 | 0.611 |
| D8 | After changing coaches or teachers, adapt to their teaching methods faster than their peers. | **0.707** | 0.438 | 0.521 |
| D14 | Often create new rules of the game with my companions. | **0.700** | 0.455 | 0.517 |
| D2 | I participate in sports activities (including sports classes and extracurricular activities) and games no less than 5 times a week. | 0.456 | **0.783** | 0.670 |
| D1 | I spend more than 2 hours on the electronic screen every day. | -0.079 | **-0.752** | 0.642 |
| D3 | Sports activities (including physical education classes and extracurricular activities) are not less than 1 hour per day. | 0.493 | **0.723** | 0.561 |
| Total Variance explained |  |  |  | 65.456 |

NOTE : Cronbach's $\alpha=0.865$ ; AC=adaptive capacity ; PAL=physical activity level.

#### 3.2.3 Results of confirmatory factor analysis(CFA)

##### 3.2.3.1 Results of first-order confirmatory factor analysis

Confirmatory factor analysis is to confirm the model of four domain structure of physical literacy on the basis of EFA, and to screen the items again. The study uses NC, RMSA, GFI, IFI, TLI, CFI to judge the overall fit of the model. The research assessed the factorial validity of the four sub-questionnaires by confirmatory factor analysis. The factor loading of all items above the standard of 0.45, ranged from 0.49 to 0.96(see Figure 3), revealing the factor validity of the measurement was satisfactory. The NC=1.859, RMSEA=0.072,GFI=0.901,IFI=0.942,TLI=0.907,GFI=0.940 (see Table 11), all indexes meet the required values, indicating that the model fits well.

**Table 11.** The first-order model fit of PLAQ

| Fit indices | NC | RMSEA | GFI | IFI | TLI | CFI |
| --- | --- | --- | --- | --- | --- | --- |
| Value | 1.859 | 0.072 | 0.901 | 0.942 | 0.907 | 0.940 |
| Discriminant value | <3 | <0.08 | >0.9 | >0.9 | >0.9 | >0.9 |

**Figure 3.**
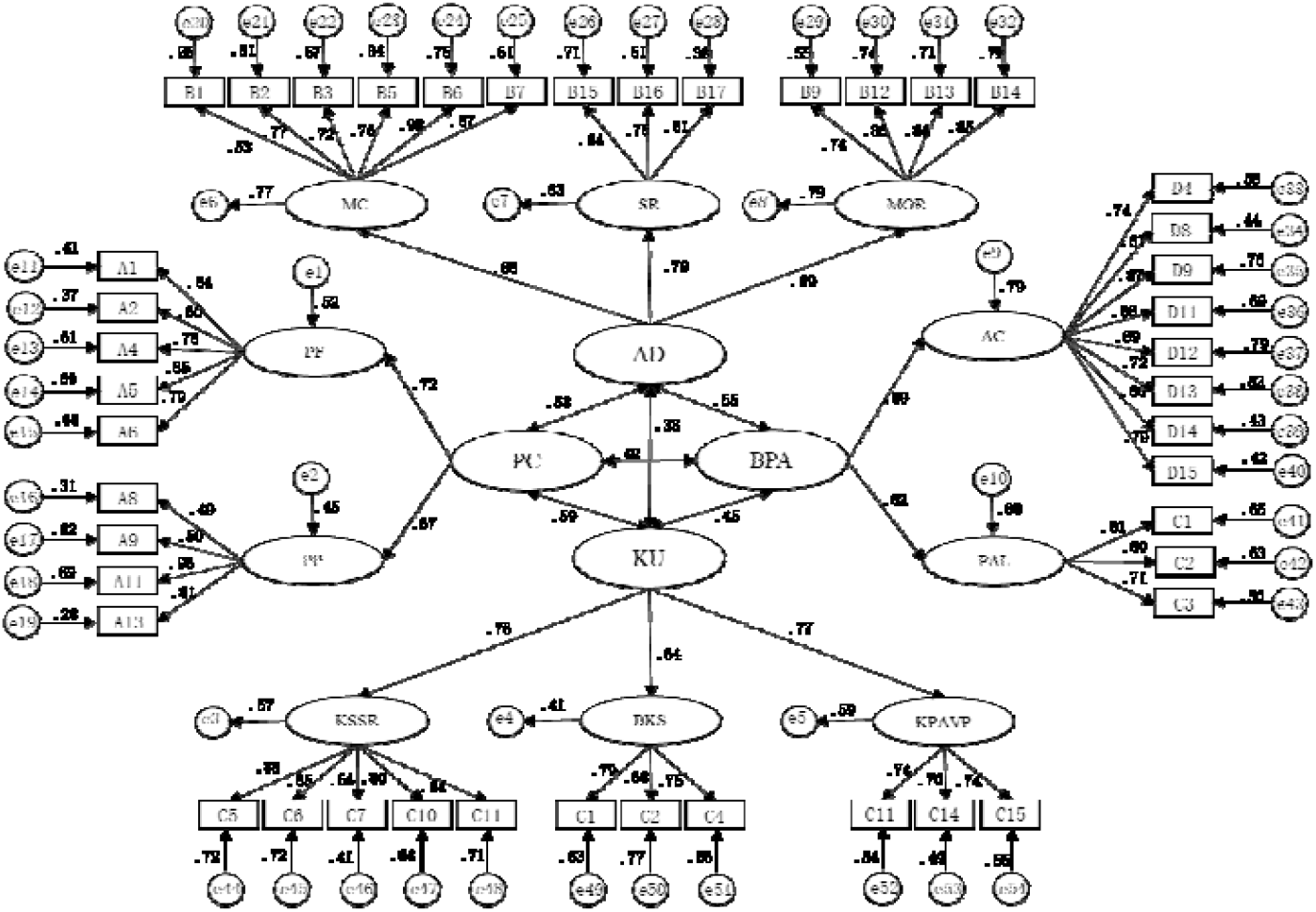
The first-order of factor structure and standardized factor loading on self- assessment physical literacy items.

For convergent validity,the AVE values of PP,PF,MC,SR,MOR,KSSR, BKS,KPAVP,AC and PAL ranged from 0.518-0.695(>0.5),and the CR values ranged from 0.770-0.923(>0.6),which indicated that the convergent validity well (see Table 12).

**Table 12.**
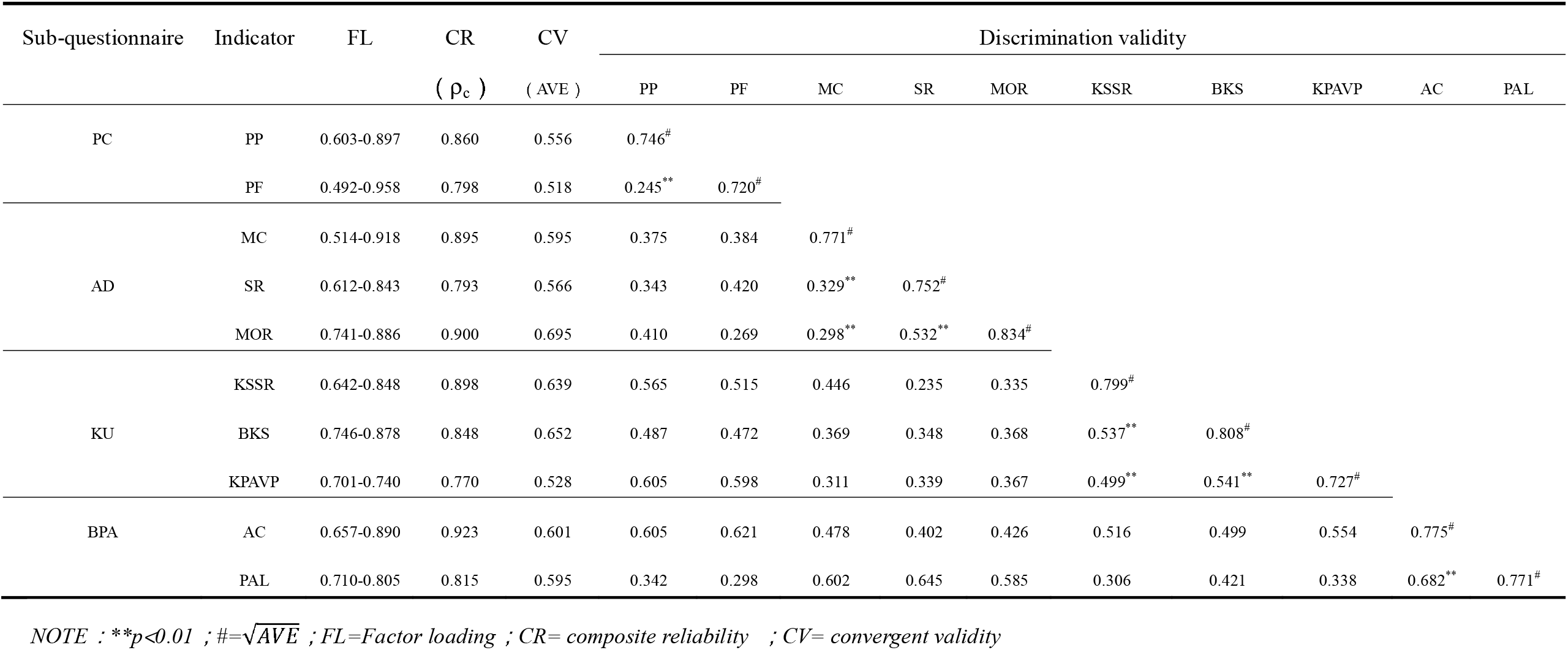
CR,CV and DV index of the first-order model fit of PLAQ

For discriminant validity, when the 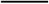 of each factor in the sub-questionnaire is higher than the value of correlation coefficient between the factors, it indicates that the factors are independent and the questionnaire has good discrimination validity. For PC sub-questionnaire,the 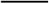 of each factors are 0.746 and 0.720,which higher than the value of correlation coefficient between the factors 0.245(see Table 12),indicating that the discriminative validity of PC sub-questionnaire is well. For AD sub-questionnaire,the 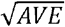 of each factors are 0.771, 0.752,0.841,which higher than the value of correlation coefficient between the factors 0.329,0.298 and 0.532,indicating that the discriminative validity of AD sub-questionnaire is well. For KU sub-questionnaire,the 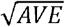 of each factors are 0.799, 0.808,0.727,which higher than the value of correlation coefficient between the factors 0.537,0.499 and 0.541,indicating that the discriminative validity of KU sub-questionnaire is well. For BPA sub-questionnaire,the 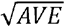 of each factors are 0.775, 0.771,which higher than the value of correlation coefficient between the factors 0.682, indicating that the discriminative validity of BPA sub-questionnaire is well.

##### 3.3.3.2 Results of Second-order confirmatory factor analysis

The second-order confirmatory factor analysis of the PLAQ uses PC, AD, KU, and BPA as latent variables. The factor loading of all items above the standard of 0.45, ranged from 0.58 to 0.92(see Figure 4), revealing the factor validity of the measurement was satisfactory. The fit indexes were all adequate as follows: NC=1.215<3, RMSEA=0.049<0.08, GFI, IFI, TLI, and CFI are all higher than 0.9 (see Table 13), which shows that the overall fitting of the model is well and can be used as an evaluation tool.

**Table 13.** The second -order model fit of PLAQ

| Fitting index | NC | RMSEA | GFI | IFI | TLI | CFI |
| --- | --- | --- | --- | --- | --- | --- |
| Value | 1.215 | 0.049 | 0.918 | 0.963 | 0.950 | 0.962 |
| Discriminant value | <3 | <0.08 | >0.9 | >0.9 | >0.9 | >0.9 |

**Figure 4.**
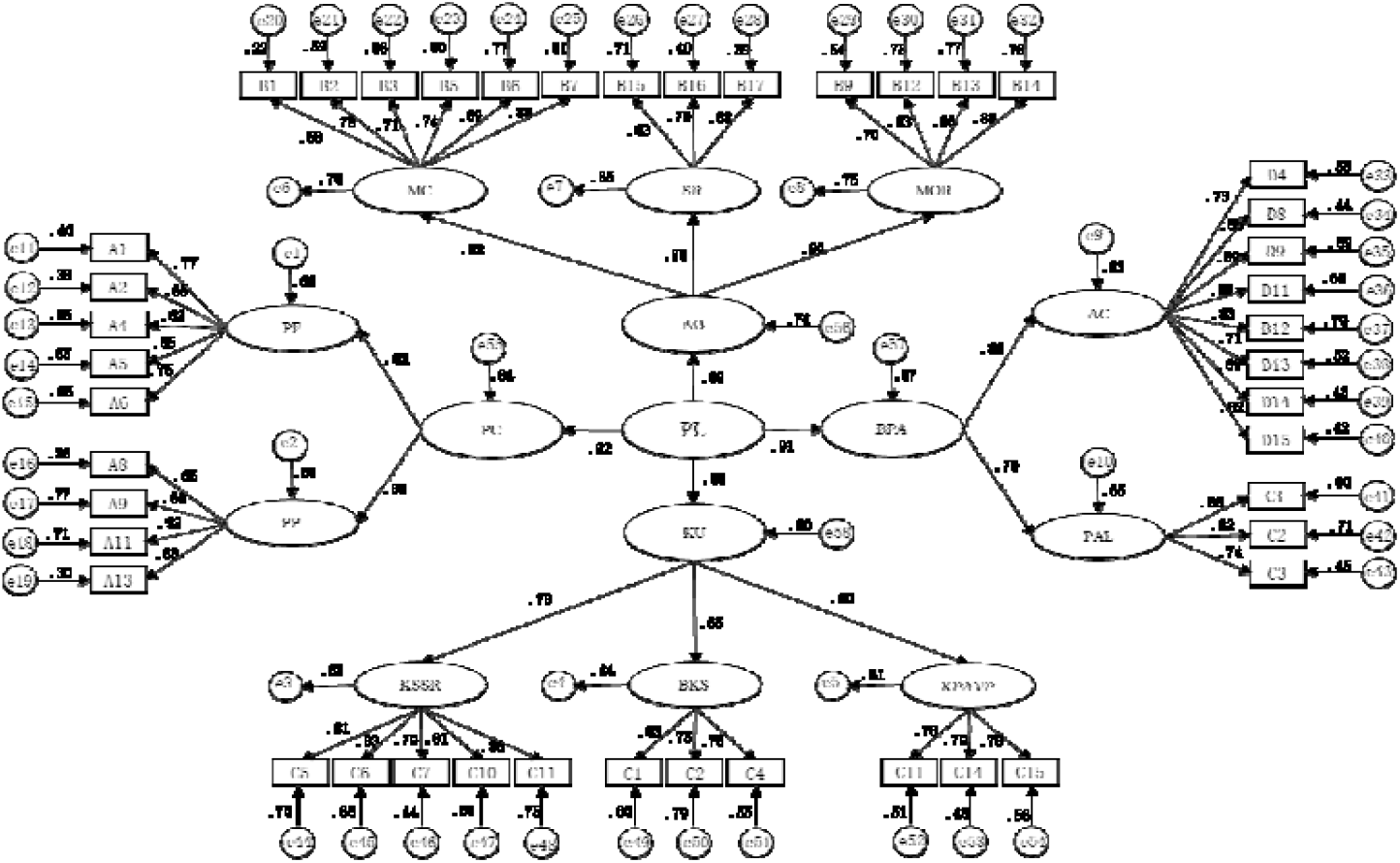
The second-order of factor structure and standardized factor loading on self- assessment physical literacy items.

## 5 Discussions

The concept, framework and measurement of PL show pluralistic characteristics in the world. Although different countries and organizations have different understandings of PL, and have proposed different versions of the definition, framework and measurement of PL, the philosophical basis of PL is highly consistent[11][25]. This diversified exploration of PL will not necessarily hinder the development of PL, but may benefit from the collision of ideas between scholars of different languages, cultures, and regions. Conversations about PL need to be more inclusive in the world.

To our knowledge,PL is a holistic and multi-dimensional concept in the world. PL is not just a certain ability related to sport, but a comprehensive trait of multiple abilities and qualities that are closely related to physical activity, which is the basis and result of physical activity. PL is a metaphor that gives name to the innate human capacity for embodied communication with the physical environment [26]. It is an ability to perceive the environment, accumulate experience, improve understanding, and use the ability to effectively interact with the world. PL is a relatively stable personality and key ability required by the continuous understanding of the body in the practice of physical activities, maintaining the individual’s continuous participation in physical activities, and using the relevant information of the body’s situation to adjust the behavior of physical activities. As stated in the definition of PL proposed by Whitehead “People with PL can perceptive in ‘reading’ all aspects of the physical environment, anticipating movement needs or possibilities and responding appropriately to these, with intelligence and imagination”[3].

The more widely used physical literacy assessment tools like CAPL,P4L and PLAY in the world generally adopt a hybrid measurement method that combines physical measurement and questionnaires,and the focus of evaluation indicators is different [27]. Owing to the evaluation standards of the above three evaluation tools are based on Canadian youths and require professional testers and facilities, the training of professional testers and the construction of facilities require a large amount of capital and human resources. Therefore, from the perspective of economy and applicability considerations are not suitable for widespread application in China. Chen Si et al. used the Delphi method to construct a Chinese adolescent physical literacy assessment tool[12], but did not report the reliability and validity of the assessment tool. Our study believed that a valid physical literacy self-assessment questionnaire (PLAQ) for Chinese primary student has yet to be developed. Compared with other assessments,we expected that the PLAQ will enable primary students, parents, teachers and other stakeholders to easily and accurately know the level of PL of primary students, thereby promoting lifelong physical activity participation.

Based on the interview data of Chinese primary students, parents, teachers and experts, this study constructed the evaluation indicators and weights applicable to the PL of Chinese students in grades 3-6. According to the evaluation indicators, a self-assessment questionnaire with good validity and reliability designed to measure for PL,and developed as an evaluation tool for the PL of students in grades 3-6. This study has a positive effect on improving the participation of Chinese adolescents in physical activity and the development of a healthy lifestyle. At the same time, it will help enrich the theoretical system of PL with Chinese cultural characteristics, and provide Chinese experience and programs for the world’s PL research.

This study proposes a four-domain structure of PL based on the Chinese population and social culture, and appeared to be consistent with international literature: physical competence, affective domain,knowledge and understanding, behavior of physical activity. The weights of the four domains of the PLAQ are different from previous PL assessments. For the PC questionnaire,PF and PP are indicators of PC that is one of the important attributes of PL. We pay more attention to students’ confidence during physical activity or exercise, and courage to compare with peers through self-perception to know their disadvantage in power,speed,endurance, coordination,flexibility,basic motor skill and specific sport skill. Good physical fitness will make children perform better in physical activity, increase confidence and promote health. International PL assessment tools also measure the physical fitness of young people from different indicators. The Chinese government proposes to promote students to master basic sports skills, improve their special skills, and enable students to master at least one sport of interest [28]. Many scholars believe that it is necessary to return to the basics and re-focus on the basic skills, required for a healthy and active life, so as to promote the formation of basic skills that required for people to participate in physical activities [10][29]. There is evidence that develop necessary skills early in life is more likely to increase physical activity throughout life and reducing the risk of chronic diseases [29]. It is vital for children to learn and apply basic skills such as running, throwing, kicking, catching and jumping with confidence and motivation.

For the AD questionnaire,we pay more attention MC,SR,and MOR indicators. The importance of MC to physical literacy has been recognized by international scholars. In addition, we further explored children’s confidence, courage and frustration during physical activity. Our research is consistent with Australia that SR is an important indicator of PL,but we designed 3 items on a five Likert-Scale for assessment and evaluation. The ability of physical and emotion self-regulation is to recognize and adjust body signals (pain, fatigue, etc.) during physical activity and adjust bad emotions is an important part of PL [29]. In traditional Chinese culture, competition and harmony are regarded as dialectical unity. Friendship, fair competition, solidify cooperation and tenacious spirit are traditional Chinese sports spirits and moral. Therefor,this study combines Chinese local sports culture and incorporates moral into the evaluation indicator of PL, in order to promote children’s attention to moral development.

In the domain of KU,the knowledge questionnaire involved in three aspects was developed,which include KSSR,BKS,and KPAVP. Edwards believes that KU are the core of cognitive ability and the key attributes of PL [11]. Knowledge guides individuals in different situations, such as what, how and when to participant in physical activity [30]. KU can change an individual’s motivation to participate in physical activity. For example, when children realize the benefits of participating in physical activity, it is possible to transform their motivation to participate in physical activity from external motivation to internal motivation. In short,KU are important domain of PL, which have a certain significance for changing physical activity motivation, developing sports skills, promoting physical activity behavior, and maintaining physical health.

In the domain of BPA,adaptive capacity and physical activity level are the main indicators that the physical activity behavior questionnaire focuses on. No matter how the PC, AD and KU changes, it will eventually be manifested in BPA. BPA is the foundation for maintaining and developing PL, and it is also the result of comprehensive application of other domains of PL. The level of physical activity is evaluated using the daily electronic screen time, daily physical activity time and weekly physical activity times. Adaptability refers to the ability to transform or change under specific conditions, it is usually a response to resiliency. Resiliency refers to the ability to respond to perceived external pressures and conflicts [31]. This study believes that when children face adversity and different situations during physical activity, they are still able to autonomous participation in physical activities as a concrete manifestation of their “reading environment”. Therefore, when compiling the questionnaire, we focus on children’s adaptive capacity to different situations.

Measurement and evaluation are the foundation to promote the development of education. If the concept of PL is to be applied in the field of education and further popularized and developed, it is necessary to developed PL assessment tools. Physical literacy is essentially a philosophical concept, and there are indicators that are difficult to understand and quantify. For example, it is difficult to evaluate the “reading environment” by using physical measurement methods. Therefore, it is necessary to build a bridge between philosophical concepts and specific evaluation indicators. This study attempts to pay attention to the situations that children face in daily life, combine the situation with evaluation indicators, and reflect a specific feature of PL through questionnaire items. The result of PLAQ is a “self-portrait” of children’s PL, and can be used as a guide for the development of PL to clarify the current status of children’s PL and the direction of future development. In addition to physical-oriented assessment, the PLAQ provides a broader assessment instrument to measure PL in this important population.

## 6 Strengths and Limitations

This study uses a combination of qualitative and quantitative research methods to construct a PLAQ for students in grades 3-6 with good reliability and validity. To our knowledge, this study is the first time that constitutes a more comprehensive interview object with students, parents, teachers and experts, and uses a bottom-up grounded theory method to construct an evaluation indicator for the PL of students in grades 3-6. When designing the items of PLAQ, the study combined local Chinese culture and focused on the real life situations faced by Chinese students in physical activities, and integrated the evaluation indicators into specific situations.

However, some limitations in this study should be mentioned. First of all, although the reliability and validity of PLAQ are good, objective measurement methods are not used to compare the results of PLAQ, so that the accuracy of PLAQ is not clear. Secondly,When constructing PL evaluation indicators according to the requirements of grounded theory, although two coders do it independently, it still cannot completely eliminate the information bias caused by the coders’ subjective factors. Thirdly,no large sample size survey have been conducted to further verify PLAQ’s effectiveness. By considering above limitations, future research should use objective measurement methods to further optimize the accuracy of PLAQ. Furthermore,in order to improve the accuracy and generalizability of PLAQ, large sample surveys for different groups may need to be conducted.

## 7. Conclusion

Under the framework of Whitehead’s PL concept, this study constructed localized 3-6 grade students’ PL evaluation indicators and assessment tools in combination with the Chinese context. Physical literacy evaluation indicators include 4 first-level indicators, 10 second-level indicators and 35 third-level indicators. In addition, the weights of indicators at all levels are also calculated, and the weight of physical activity behavior is the largest, which is different from the western physical competence orientation. This study developed the PLAQ for Chinese students in grades 3-6 that are grouped within four primary domains:physical competence(PC), affective domain(AD),knowledge and understanding(KU),behavior of physical activity(BPA). The PLAQ is composed of 44 items, and the reliability and validity of the questionnaire is well, which can be used as an evaluation tool for the PL of stakeholders. Although this study constructed the model of the PL structure and developed the PL assessment tool for Chinese students in grades 3-6, the accuracy of the tool needs to be further verified. Therefore, in the future, a large-scale survey is needed to compare and optimize the accuracy and generalizability of PL assessment tools.

## Data Availability

All relevant data are within the manuscript and its Supporting Information files.

## Notes

### Competing Interest Statement

The authors have declared no competing interest.

### Funding Statement

The author(s) received no specific funding for this work.

### Author Declarations

Academic Committee of the School of Physical Education and Sport Science, Fujian Normal University gave ethical approval for this work.

